# Cytotoxic lymphocytes are dysregulated in multisystem inflammatory syndrome in children

**DOI:** 10.1101/2020.08.29.20182899

**Authors:** Noam D. Beckmann, Phillip H. Comella, Esther Cheng, Lauren Lepow, Aviva G. Beckmann, Konstantinos Mouskas, Nicole W. Simons, Gabriel E. Hoffman, Nancy J. Francoeur, Diane Marie Del Valle, Gurpawan Kang, Emily Moya, Lillian Wilkins, Jessica Le Berichel, Christie Chang, Robert Marvin, Sharlene Calorossi, Alona Lansky, Laura Walker, Nancy Yi, Alex Yu, Matthew Hartnett, Melody Eaton, Sandra Hatem, Hajra Jamal, Alara Akyatan, Alexandra Tabachnikova, Lora E. Liharska, Liam Cotter, Brian Fennessey, Akhil Vaid, Guillermo Barturen, Scott R. Tyler, Hardik Shah, Ying-chih Wang, Shwetha Hara Sridhar, Juan Soto, Swaroop Bose, Kent Madrid, Ethan Ellis, Elyze Merzier, Konstantinos Vlachos, Nataly Fishman, Manying Tin, Melissa Smith, Hui Xie, Manishkumar Patel, Kimberly Argueta, Jocelyn Harris, Neha Karekar, Craig Batchelor, Jose Lacunza, Mahlet Yishak, Kevin Tuballes, Leisha Scott, Arvind Kumar, Suraj Jaladanki, Ryan Thompson, Evan Clark, Bojan Losic, The Mount Sinai COVID-19 Biobank Team, Jun Zhu, Wenhui Wang, Andrew Kasarskis, Benjamin S. Glicksberg, Girish Nadkarni, Dusan Bogunovic, Cordelia Elaiho, Sandeep Gangadharan, George Ofori-Amanfo, Kasey Alesso-Carra, Kenan Onel, Karen M. Wilson, Carmen Argmann, Marta E. Alarcón-Riquelme, Thomas U. Marron, Adeeb Rahman, Seunghee Kim-Schulze, Sacha Gnjatic, Bruce D. Gelb, Miriam Merad, Robert Sebra, Eric E. Schadt, Alexander W. Charney

## Abstract

Multisystem inflammatory syndrome in children (MIS-C) presents with fever, inflammation and multiple organ involvement in individuals under 21 years following severe acute respiratory syndrome coronavirus 2 (SARS-CoV-2) infection. To identify genes, pathways and cell types driving MIS-C, we sequenced the blood transcriptomes of MIS-C cases, pediatric cases of coronavirus disease 2019, and healthy controls. We define a MIS-C transcriptional signature partially shared with the transcriptional response to SARS-CoV-2 infection and with the signature of Kawasaki disease, a clinically similar condition. By projecting the MIS-C signature onto a co-expression network, we identified disease gene modules and found genes downregulated in MIS-C clustered in a module enriched for the transcriptional signatures of exhausted CD8^+^ T-cells and CD56^dim^CD57^+^ NK cells. Bayesian network analyses revealed nine key regulators of this module, including *TBX21*, a central coordinator of exhausted CD8^+^ T-cell differentiation. Together, these findings suggest dysregulated cytotoxic lymphocyte response to SARS-Cov-2 infection in MIS-C.

## Introduction

Multisystem inflammatory syndrome in children (MIS-C) is a novel condition thought to be secondary to severe acute respiratory syndrome coronavirus 2 (SARS-CoV-2) infection(Dufort et al., 2020; Riphagen et al., 2020). Clinically distinct from coronavirus disease 2019 (COVID-19), MIS-C presents in individuals below the age of 21 as fever, inflammation, severe involvement of at least two organ systems (cardiac, renal, respiratory, hematologic, gastrointestinal, dermatologic or neurological systems), and current or recent SARS-CoV-2 infection or exposure(2020). At the time of writing, approximately 600 cases of MIS-C have been reported in the United States, the majority of which had no underlying medical conditions(Godfred-Cato, 2020). These numbers are expected to increase with the number of pediatric COVID-19 cases(American Academy of Pediatrics and the Children’s Hospital Association, 2020) as the disease is reported to arise ~6 weeks after SARS-CoV-2 infection(Gruber et al., 2020). MIS-C shares symptoms with Kawasaki disease (KD) and other pediatric inflammatory conditions, and was initially termed the “Kawasaki-like disease”(Verdoni et al., 2020). With the genes and cell types involved in the disease unknown, its similarity to KD at the molecular level has not been established, and while hypotheses have been put forth that it is a systemic autoimmune disease triggered by SARS-CoV-2 infection(Cavounidis et al., 2020; Consiglio et al.; Gruber et al., 2020), its pathogenesis has yet to be definitively determined.

Here, we report the results of a genome-wide study, mapped out in Figure 1, of the molecular architecture of MIS-C. RNA sequencing (RNA-seq) was generated on 30 whole blood samples from MIS-C cases (8 specimens from 8 individuals), pediatric COVID-19 cases (18 specimens from 7 individuals), and healthy controls (HCs; 4 specimens from 4 individuals) *(Figure 1A)*. Three primary analyses were performed on this data: cell type deconvolution *(Figure 1B)*, differential expression (DE, *Figure 1C)*, and co-expression network construction *(Figure 1D)*. Deconvolution estimated the relative cell type proportions in each transcriptome, which were then compared between cases and controls to identify the immune cell types involved in the pathogenesis of MIS-C *(Figure 1B)*. Expression of each gene in the transcriptome was tested for association with disease, resulting in a MIS-C signature that was then queried to resolve the dysregulated molecular pathways *(Figure 1C)*. Co-expression network construction organized genes into coherent units called modules *(Figure 1D)* and the modules loaded with genes of the MIS-C signature were empirically identified *(Figure 1E)*. These modules were functionally annotated to pathway and cell type signatures, and we explored their role in other diseases of interest *(Figure 1F)*. The module with the strongest enrichment for MIS-C was further annotated to pinpoint cell subtypes, and key regulators of the processes captured by this module were identified in a regulatory network built from whole blood gene expression in a large cohort of individuals with inflammatory bowel disease (IBD, *Figure 1G*). This stepwise approach shed light on several questions regarding the underpinnings of MIS-C. It revealed a partially shared molecular etiology with KD but not other pediatric inflammatory conditions, no overlap with the transcriptional signatures of classic autoimmune diseases, and a direct link with the immune response to SARS-CoV-2 infection. Evidence converged on the downregulation of exhausted CD8^+^ T-cells and CD56^dim^CD57^+^ natural killer (NK) cells, with *TBX21, TGFBR3*, and 7 other genes the key drivers of MIS-C pathogenesis.

**Figure 1:**
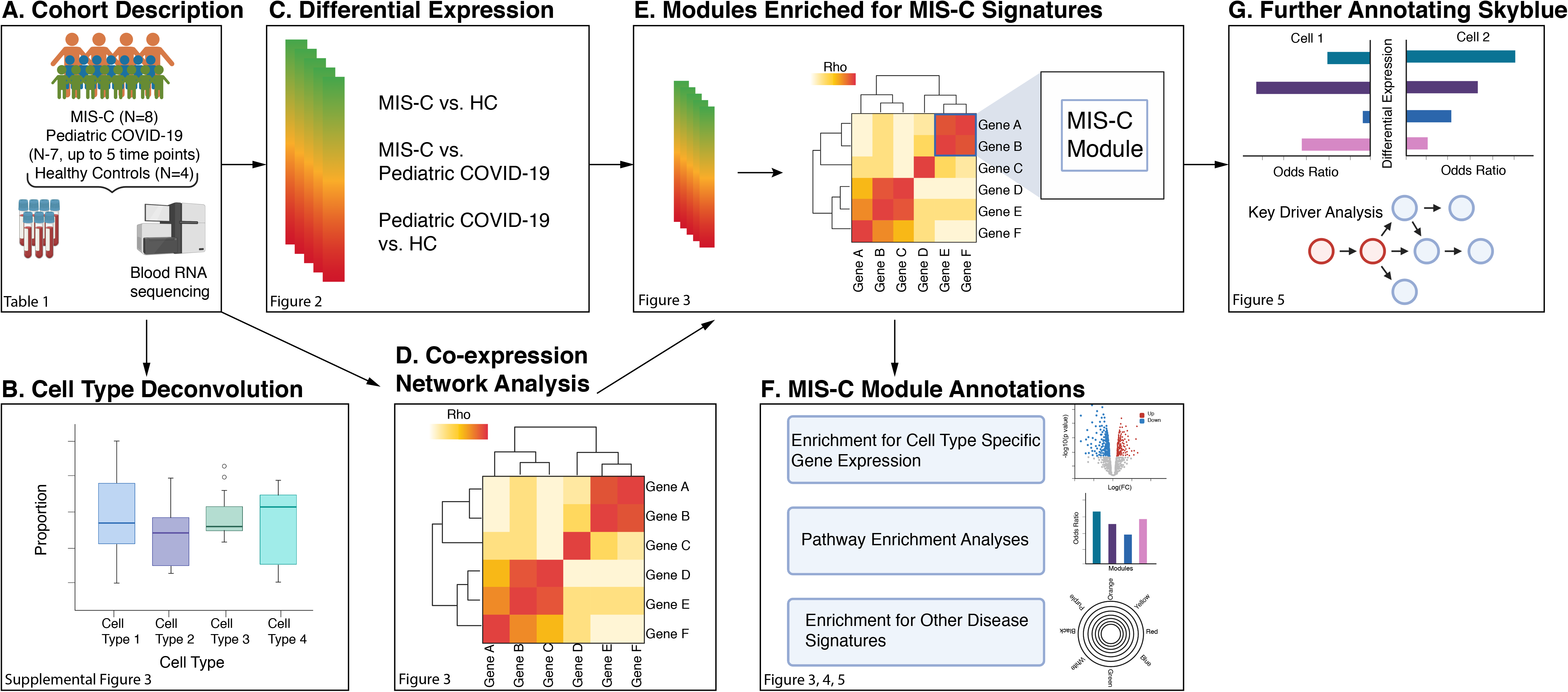
Study Overview. Workflow of the analyses presented in the paper.

## Results

### Cohort description

MIS-C cases, pediatric COVID-19 cases, and HCs (N=8, 7, and 4, respectively) were recruited with case referrals coming from treating physicians. Inclusion criteria were age less than 21 years and fulfillment of the formal MIS-C diagnostic criteria(2020) (for MIS-C cases) or a recent diagnosis of COVID-19 in the absence of suspicion for MIS-C (for pediatric COVID-19 cases). MIS-C patients ranged in age from five to 20 years old (mean = 11, standard deviation = 5.2) and were largely free of other comorbidities. Nearly all of the seven pediatric COVID-19 cases (mean age = 12.4, standard deviation = 7) were chronically immunocompromised, with three suffering from comorbid malignancies requiring chemotherapy and three on immunosuppressive therapy for other indications. Recruitment of age-matched HCs was not feasible during the study period due to the risk of exposing healthy children to COVID-19 by coming to the hospital for a research blood draw. HCs for this study therefore consisted of healthy adults working on-site during the study period (N = 4, mean age = 30.75, standard deviation = 6.18). Key demographic and clinical variables for all participants are detailed in *Table 1* and *Data S1*. Informed consent was obtained from either the participant or the participant’s legally authorized representative, and all study-related activities were conducted under the approval and oversight of Mount Sinai’s Institutional Review Board.

**Table 1:**
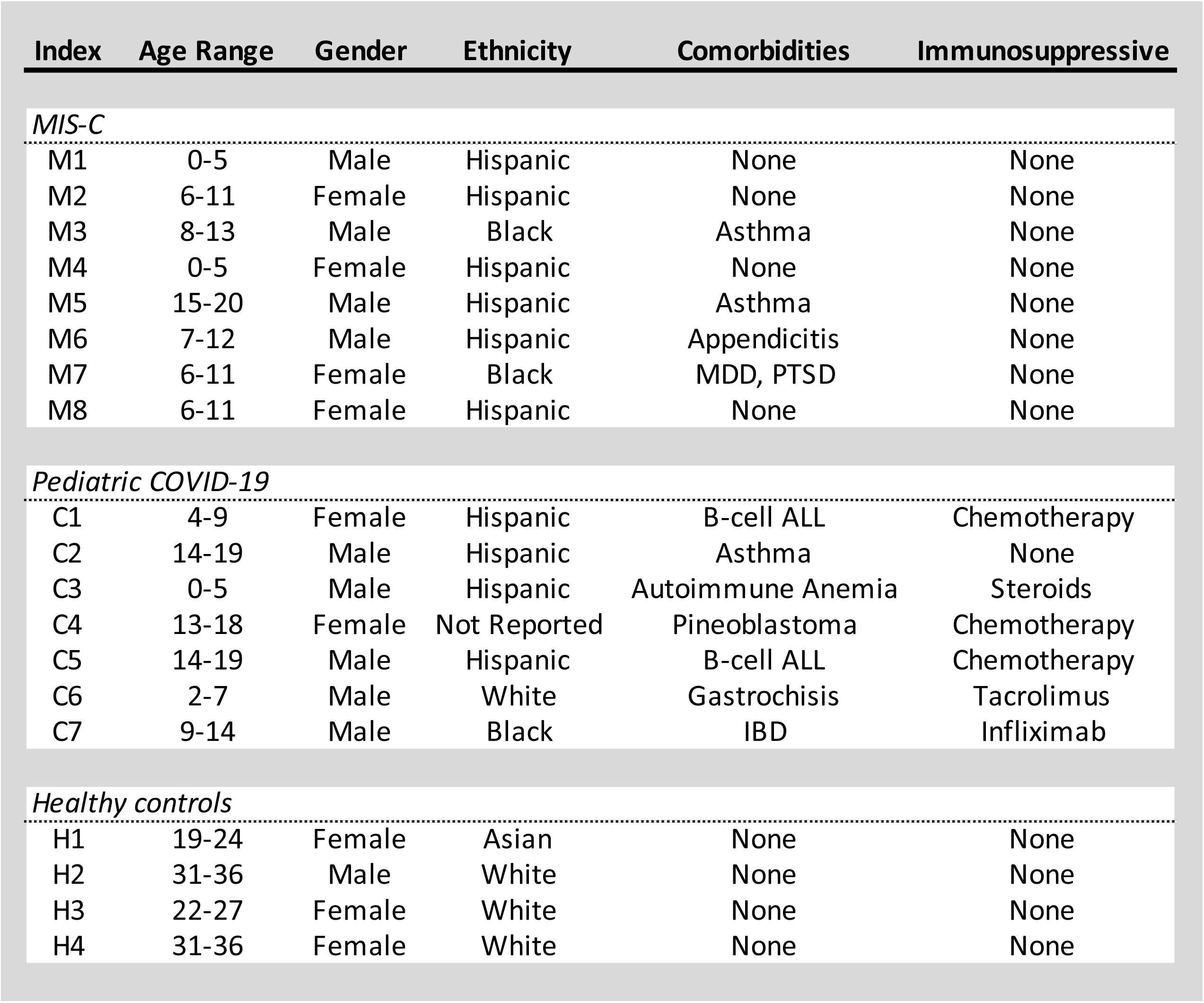
Sample Description. Demographic and clinical variables for the MIS-C cases (N = 8), pediatric COVID-19 cases (N = 7), and healthy controls (N = 4). Index column refers to the individual identifiers. Age is presented in years. Sex, ethnicity, and comorbidities are as reported in the electronic medical record. Immunosuppressants indicate the usage of chronic immunosuppressive therapies for any indication. Further clinical data can be found in ***Data S1***. MDD = Major Depressive Disorder; PTSD = Post-Traumatic Stress Disorder; ALL = Acute Lymphoblastic Leukemia, IBD = Inflammatory Bowel Disease.

| Index | Age Range | Gender | Ethnicity | Comorbidities | Immunosuppressive |
| --- | --- | --- | --- | --- | --- |
| *MIS-C* |  |  |  |  |  |
| M1 | 0-5 | Male | Hispanic | None | None |
| M2 | 6-11 | Female | Hispanic | None | None |
| M3 | 8-13 | Male | Black | Asthma | None |
| M4 | 0-5 | Female | Hispanic | None | None |
| M5 | 15-20 | Male | Hispanic | Asthma | None |
| M6 | 7-12 | Male | Hispanic | Appendicitis | None |
| M7 | 6-11 | Female | Black | MDD, PTSD | None |
| M8 | 6-11 | Female | Hispanic | None | None |
| *Pediatric COVID-19* |  |  |  |  |  |
| C1 | 4-9 | Female | Hispanic | B-cell ALL | Chemotherapy |
| C2 | 14-19 | Male | Hispanic | Asthma | None |
| C3 | 0-5 | Male | Hispanic | Autoimmune Anemia | Steroids |
| C4 | 13-18 | Female | Not Reported | Pineoblastoma | Chemotherapy |
| C5 | 14-19 | Male | Hispanic | B-cell ALL | Chemotherapy |
| C6 | 2-7 | Male | White | Gastrochisis | Tacrolimus |
| C7 | 9-14 | Male | Black | IBD | Infliximab |
| *Healthy controls* |  |  |  |  |  |
| H1 | 19-24 | Female | Asian | None | None |
| H2 | 31-36 | Male | White | None | None |
| H3 | 22-27 | Female | White | None | None |
| H4 | 31-36 | Female | White | None | None |

### Quality control and accounting for confounding effects of age

RNA-seq data is often impacted by unwanted technical and biological variation that needs to be accounted for prior to any analysis. We explored technical and biological drivers of variance using a combination of principal component and variancePartition analyses(Hoffman and Roussos, 2020; Hoffman and Schadt, 2016) on the normalized count data. These analyses identified a set of technical and biological covariates that explained a substantial fraction of the expression variation in the dataset and were subsequently accounted for using linear mixed modeling in the differential expression (DE) and co-expression network analyses performed *(Figure S1A, B, C and* D).

Age is known to contribute to variation in gene expression(Solana et al., 2012) but in this study recruitment of age-matched HCs was impractical for reasons stated above, leading to a strong correlation between age and disease status *(Figure S1B)*. Leveraging published age gene DE signatures from whole blood(Lin et al., 2019; Peters et al., 2015; Yang et al., 2015), we identified 23 modules in a co-expression network built from our data after covariate adjustment significantly enriched for genes associated with age (see *Methods, Data S2-2)*. To account for this age effect on the gene expression variation without masking the true MIS-C disease signature, we generated a metric for each individual in our cohort to capture the transcriptional effects of age. Using the co-expression network defined above, we extracted all genes in any module significantly enriched for age DE signatures and generated a cross-module eigengene value (Spearman’s rho for the correlation of this metric with age = -0.32, p-value = 0.047). We observed that it had an effect on the general patterns of expression in our data without confounding disease state, as illustrated by principal components analyses before and after inclusion of imputed age in the model (respectively *Figures S2A and B)*. This eigengene was included as an additional covariate in all DE and co-expression network analyses performed below *(Figure S1C, D, S2)*.

### Cell type deconvolution implicates CD8^+^ T-cells in MIS-C

We utilized Cibersortx(Newman et al., 2019) to deconvolve gene expression for each sample in our dataset into composite blood cell fractions *(Figure 1B)*. Three sources of peripheral blood mononuclear cell (PBMC) reference transcriptomes were used, one derived from collection of transcriptomic profiles from the public domain (the “LM22” dataset) and two from single-cell RNA-seq (the “NSCLC” and “SCP424” datasets). In MIS-C cases compared to controls, downregulation of CD8^+^ T-cells was observed in LM22 (mean relative cell proportion change beta = -0.09, adjusted p-value = 0.04) and SCP424 (beta = -0.13, adjusted p-value = 0.01), while downregulation of NK T-cells was observed in NSCLC (beta = -0.05; adjusted p-value = 0.01). B-cell signatures in pediatric COVID-19 cases were downregulated compared to MIS-C cases (B-cell signature in SCP424 beta = -0.11, adjusted p-value = 1.13E10-5; plasma cell signature in LM22 beta = -0.01, adjusted p-value = 0.046) and upregulated compared to controls (B-cell signature in SC424 beta = 0.06, adjusted p-value = 0.04) *(Figure S3; Data S3)*.

To validate these findings, we correlated estimated cell type fractions with complete blood counts (CBCs) performed by the clinical laboratory as standard-of-care during hospitalization. Significant correlations were observed between the CBC result and LM22 estimates for neutrophils (Spearman’s rho = 0.83, adjusted p-value = 0.011) and monocytes (Spearman’s rho = 0.86, adjusted p-value = 0.007). To compare lymphocytes from the CBCs with deconvolution estimates, we derived a single lymphocyte estimate for each of the three references used by summing all T- and B-cell type fractions (see *Methods)*. The deconvolution-derived estimates for lymphocytes were significantly correlated with the CBC for all three references (LM22 Spearman’s rho = 0.77, adjusted p-value = 0.015; SCP424 Spearman’s rho = 0.78, adjusted p-value = 0.028; NSCLC Spearman’s rho = 0.89, adjusted p-value = 0.003) (*Figure S3D)*.

### Defining a MIS-C differential expression signature

To gain insight into the molecular pathology of MIS-C, we performed DE analyses comparing each phenotype pair in our dataset *(Figure 1C)*. In order to retain power while accurately controlling the false positive rate it was essential to account for the repeated measures in pediatric COVID-19 cases and the technical replicates in HCs (see *Methods)*. Towards this end, we used dream(Hoffman and Roussos, 2020), which models the effect of the individual on gene expression as a random effect for each gene using a linear mixed model. We ran three DE analyses in our cohort: MIS-C cases compared to HCs, pediatric COVID-19 cases compared to HCs, and MIS-C compared to pediatric COVID-19 cases *(Data S4-1)*. Multiple test correction was performed separately for each comparison, accounting for the number of genes tested using the method of Benjamini and Hochberg(Hochberg and Benjamini, 1990).

Few differentially expressed genes (DEGs) were observed after multiple test correction (8 DEGs for pediatric COVID-19 versus HC, 1 DEG for pediatric COVID-19 versus MIS-C), but we identified many DEGs with unadjusted p-value < 0.05 *(Figure 2A, Figure 2B; Data S4-1)*. This result is not itself surprising given the small sample size, our efforts to avoid false positives by accounting for 8 biological and technical covariates, and inclusion of the predicted age effect on gene expression in order to protect against age being confounded with disease status. Sub-significant gene expression signatures can still capture substantial biological signals in the ranking of genes(Storey and Tibshirani, 2003; Subramanian et al., 2005; Wu and Smyth, 2012). To estimate whether there was evidence of true signal associated to disease despite the lack of genome-wide significant DEGs, we estimated the tt_1_ value, which provides a lower bound of the proportion of genes tested that truly deviate from the null hypothesis(Storey and Tibshirani, 2003) and found that approximately 15% of the genes tested between MIS-C and HCs likely are true DEGs. We therefore defined MIS-C DEGs as the set of 2,043 genes (815 upregulated and 1,228 downregulated) with an unadjusted p-value below 0.05 (hereafter, the “MIS-C signature”). We also defined a pediatric COVID-19 signature (2,505 genes; 1,232 upregulated and 1,273 downregulated) and a signature differentiating MIS-C from pediatric COVID-19 (2,062 genes; 1,295 upregulated in MIS-C and 767 upregulated in pediatric COVID-19) *(Data S4-1)*.

To identify known biological processes associated with MIS-C and pediatric COVID-19, we performed pathway enrichment analyses on the three upregulated and downregulated DE signatures *(Data S4-2 and Figure 2C;* see *Methods)*. The top two gene ontology (GO) terms associated to the upregulated MIS-C signature were ‘regulated exocytosis’ (odds ratio, OR=3.23, false discovery rate, FDR = 1.07E-20) and ‘secretion by cell’ (OR=2.54, FDR=4.19E-20E-19). Other enriched GO terms were either related to innate immunity or non-specific references to the immune system. Downregulated genes in MIS-C were enriched for ‘RNA metabolic process’ (OR=1.41, FDR=4.34E-11), ‘nucleic acid metabolic process’ (OR=1.36, FDR=3.25E-10), and other GO terms related to the regulation of gene expression. The upregulated pediatric COVID-19 DE signature was enriched for the GO terms ‘vesicle-mediated transport’ (OR=1.61, FDR=2.17E-09), ‘cellular catabolic process’ (OR=1.53, FDR=3.83E-08), and ‘catabolic process’ (OR=1.50, FDR=5.48E-08) and the downregulated signature was enriched for ‘regulation of transcription by RNA polymerase’ (OR=1.5, FDR=1.82e-06) and ‘regulation of RNA biosynthetic process’ (OR=1.38, FDR=1.57E-05). Comparing MIS-C to pediatric COVID-19, genes upregulated in MIS-C were enriched for ‘immune response’ (OR=1.66, FDR=0.0028) and ‘immune system process’ (OR=1.51, FDR=2.81E-03) while genes upregulated in pediatric COVID-19 were enriched for ‘cellular macromolecule catabolic process’ (OR=1.88, FDR=3.79e-11) and ‘cellular protein catabolic process’ (OR=2.04, FDR=1.30E-09) *(Data S4-2)*.

**Figure 2:**
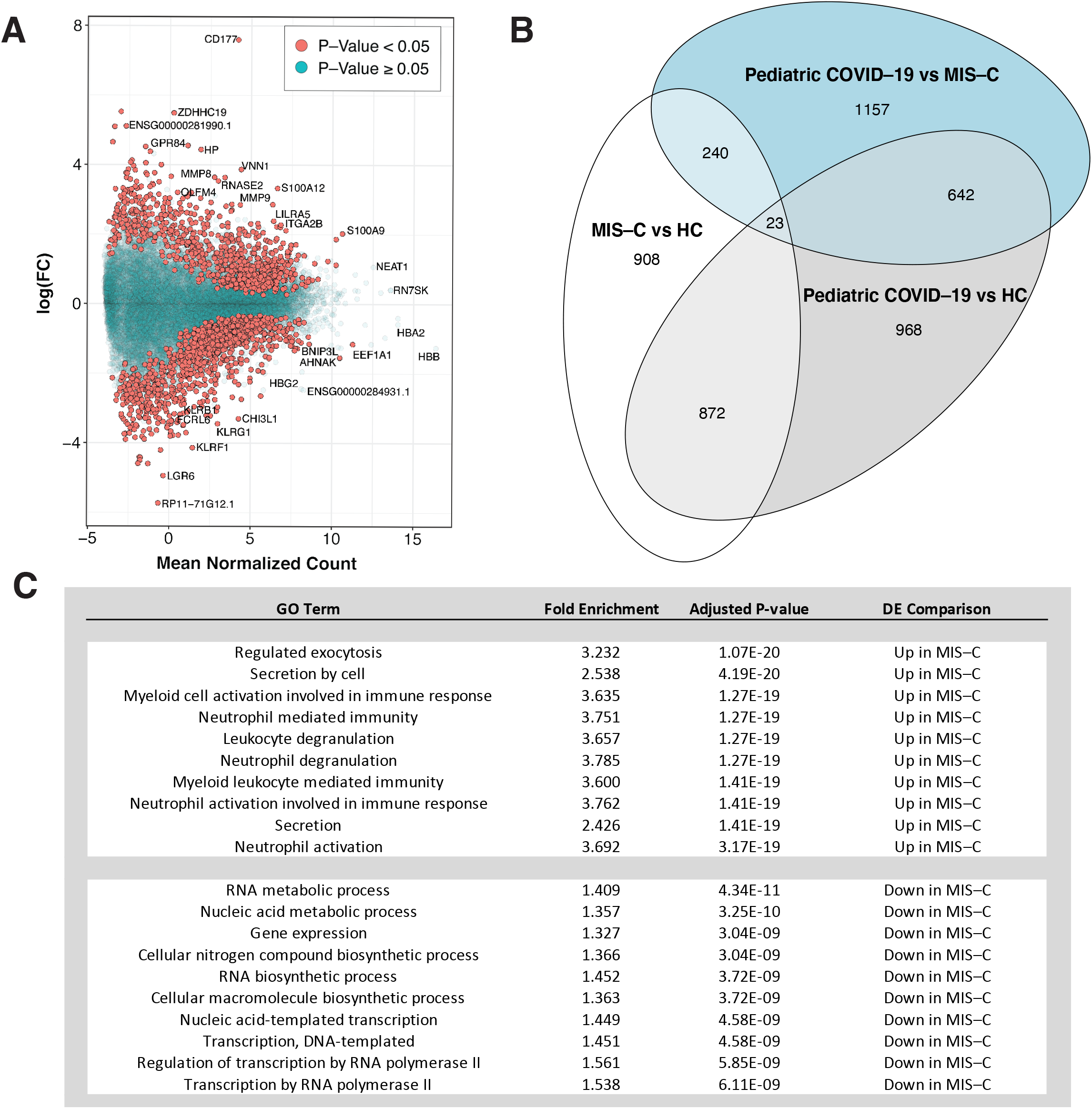
Differential expression analyses identify the transcriptional signature of MIS-C. A: Differential expression (DE) analyses for MIS-C patients versus HCs. The x-axis is the mean normalized count for each gene and the y-axis the log_2_(fold-change) for the differential expression. Positive and negative log_2_(fold change) represent genes upregulated and downregulated in MIS-C, respectively, and significance of association between gene expression and MIS-C status is indicated by the color of the dots as defined in the legend. B: Overlap of MIS-C and pediatric COVID-19 transcriptional signatures: Venn diagram of the overlap of genes across DE signatures. Each comparison is labeled on the plot. C: GO terms for MIS-C signature: GO term enrichment results for the top 10 upregulated and downregulated processes in MIS-C compared to HCs. Full DE results and pathway enrichments for all comparisons in B can be found in ***Data S4***.

### Dissecting the MIS-C signature with co-expression network analyses

DE analyses permit the broad detection of genes and pathways associated with disease states. Co-expression network analysis builds on this by leveraging the correlation across the transcriptome to organize genes into functional units called modules, sometimes linked to specific cell types and biological processes(Langfelder et al., 2013). To further dissect the MIS-C signature, weighted gene co-expression network analysis (WGCNA)(Langfelder and Horvath, 2008) was used to construct a network comprising 37 non-overlapping modules *(Figure 1D)*.

To maximize the interpretability of coexpression networks, modules can be annotated to the biological pathways, cell types, and disease processes they capture. Annotation in this context consists of projecting a feature set of interest onto the network and performing an enrichment test for the overlap between the features in the set and the genes in the module. Feature sets tested here included gene sets from pathway databases, cell type markers, and DEGs from our DE analyses and the literature (see *Methods; Data S2-3,5,6,7, Figure 1E, Figure 3*, S4)(Ashburner et al., 2000; Subramanian et al., 2005). Many modules were significantly associated with GO terms *(Figure 3A, Data S2-5)*, MSigDB Hallmark terms *(Data S2-6)*, and MSigDB C7 terms *(Data S2-7)* that were consistent with their cell type annotations. As an example, the darkolivegreen module was consistently enriched for B-cell signatures (adjusted p-value < 0.05 in 6 B-cell signatures from multiple independent references; *Figure 3B, S4;* see *Methods)*. Other modules were enriched for multiple cell type signatures and/or pathways. The salmon module, for instance, was enriched for signatures of B-cells, monocytes, and dendritic cells (adjusted p-values < 0.05), as well as for the GO term ‘MHC class II protein’ (OR=27.03, FDR=2.8E-09). Taken together, these annotations suggested a biologically coherent network structure, and this annotated co-expression network therefore formed the basis for much of the remaining analyses in this report. We next projected the upregulated and downregulated MIS-C signatures onto the network to identify the modules with more DEGs than would be expected by chance. After accounting for multiple testing (see *Methods)*, 11 of the 37 modules were enriched for MIS-C DEGs, with 6 modules enriched for upregulated genes, 9 modules enriched for downregulated genes, and 4 modules enriched for both. (*Figure 3C; see Methods*).

**Figure 3:**
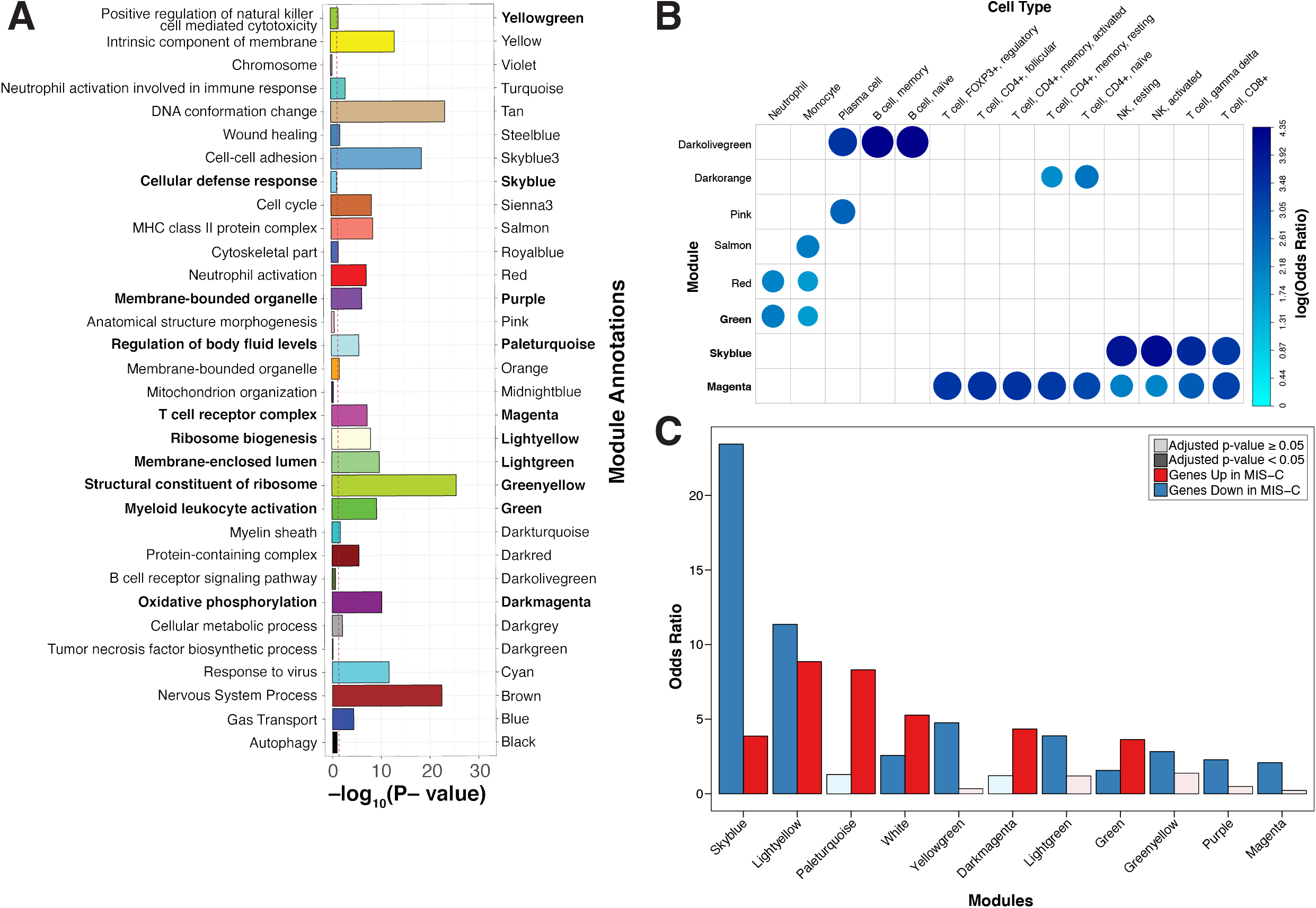
Co-expression network analysis identifies modules of genes dysregulated in MIS-C. A: Module GO term enrichments. The x-axis is the most significant GO term associated with each module, the y-axis is the -log_10_(adjusted p-value) for the enrichment. Bars are colored by module names, which are also specified for clarity. Only modules with an enrichment p-value < 1 are shown. The threshold for significance threshold, -log_10_(0.05), is indicated by the red dashed line. In bold lettering are the modules enriched for the MIS-C signature. B: Module cell type signature enrichments. The x-axis and y-axis are the names of the modules and of the cell type signatures, respectively. All signatures shown here are derived from the LM22 reference (Newman et al, *Nature Methods*, 2015). The color and size of the circles represents the log_2_(odds ratio) of the enrichments as defined in the legend. In bold are the modules enriched for the MIS-C signature. C: Modules enrichment for MIS-C signatures. The x-axis is the module names and the y-axis the odds ratio of the enrichment of the modules for the genes upregulated and downregulated in MIS-C. Only modules significantly enriched for MIS-C DEGs are shown. The color of the bars represent direction and the opacity represent the significance as defined in the legend. All module enrichments can be found in ***Data S2***.

The clinical similarity between MIS-C and KD has led to the suggestion they have molecular similarities(Verdoni et al., 2020). We assessed this by testing whether the modules enriched for MIS-C DEGs were also enriched for KD DEGs. KD DEGs were generated by re-analysis of publicly available whole blood transcriptomic data from 78 KD patients and 55 controls(Wright et al., 2018) (see *Methods)*. We observed that 4 modules were enriched for both MIS-C and KD in the same direction, indicating partially shared molecular etiology and/or common downstream effects between these traits *(Figure 4, Data S2-4)*. The module most significantly impacted by MIS-C and KD was the skyblue module, comprised of 159 genes, which was significantly enriched for downregulated DEGs in both MIS-C (Fisher’s exact test odds ratio, OR=23.44, adjusted p-value=4.33E-66) and KD (Fisher’s exact test OR=19.18, adjusted p-value=1.64E-04). This module was annotated to CD8^+^ T-cells and NK cells, both essential components of the cytotoxic immune response *(Figure 3B, Data S2-3; see Methods)*. The magenta module was also enriched for downregulated DEGs in MIS-C (Fisher’s exact test OR=2.08, adjusted p-value=2.81E-04) and KD (Fisher’s exact test OR=12.62, adjusted p-value=4.29E-08) and also annotated for CD8^+^ T-cells and NK cells *(Figure 3B, Data S2-3; see Methods)*. Modules that were enriched for either MIS-C or KD but not both included paleturquoise, which had the largest unique enrichment for upregulated DEGs in MIS-C (Fisher’s exact test OR=8.31, adjusted p-value=4.12E-15), but not KD (Fisher’s exact test OR=2.43, adjusted p-value=1.0), and the red module, which was enriched for upregulated DEGs in KD (Fisher’s exact test OR=2.65, adjusted p-value=3.45E-10) but not MIS-C (Fisher’s exact test OR=0.28, adjusted p-value=1.0). Paleturquoise was enriched for genes expressed in platelets (maximum Fisher’s exact test OR=86.09, adjusted p-value=6.37E-15) and the GO term ‘coagulation activity’ (OR=8.02, FDR=3.04E-06), while the red module was annotated to myeloid cell types such as mononuclear phagocytes and neutrophils and GO pathway ‘neutrophil activation’ (OR=2.32, FDR=6.5E-08) *(Figure 3B, Data S2-5; see Methods)*. To assess whether this overlap with MIS-C was unique to KD or shared with other multisystem inflammatory conditions seen in pediatric populations, we projected signatures for Macrophage Activation Syndrome (MAS) and Neonatal-Onset Multisystem Inflammatory Disease (NOMID) onto the network(Canna et al., 2014). In contrast to KD, minimal overlap was seen between these conditions and MIS-C, with only one module enrichment shared between MIS-C and MAS (for upregulated DEGs in the green module; *Figure 4)* and none between MIS-C and NOMID. Altogether, these observations support shared molecular etiology and/or common downstream effects between KD and MIS-C in addition to their clinical similarities.

**Figure 4:**
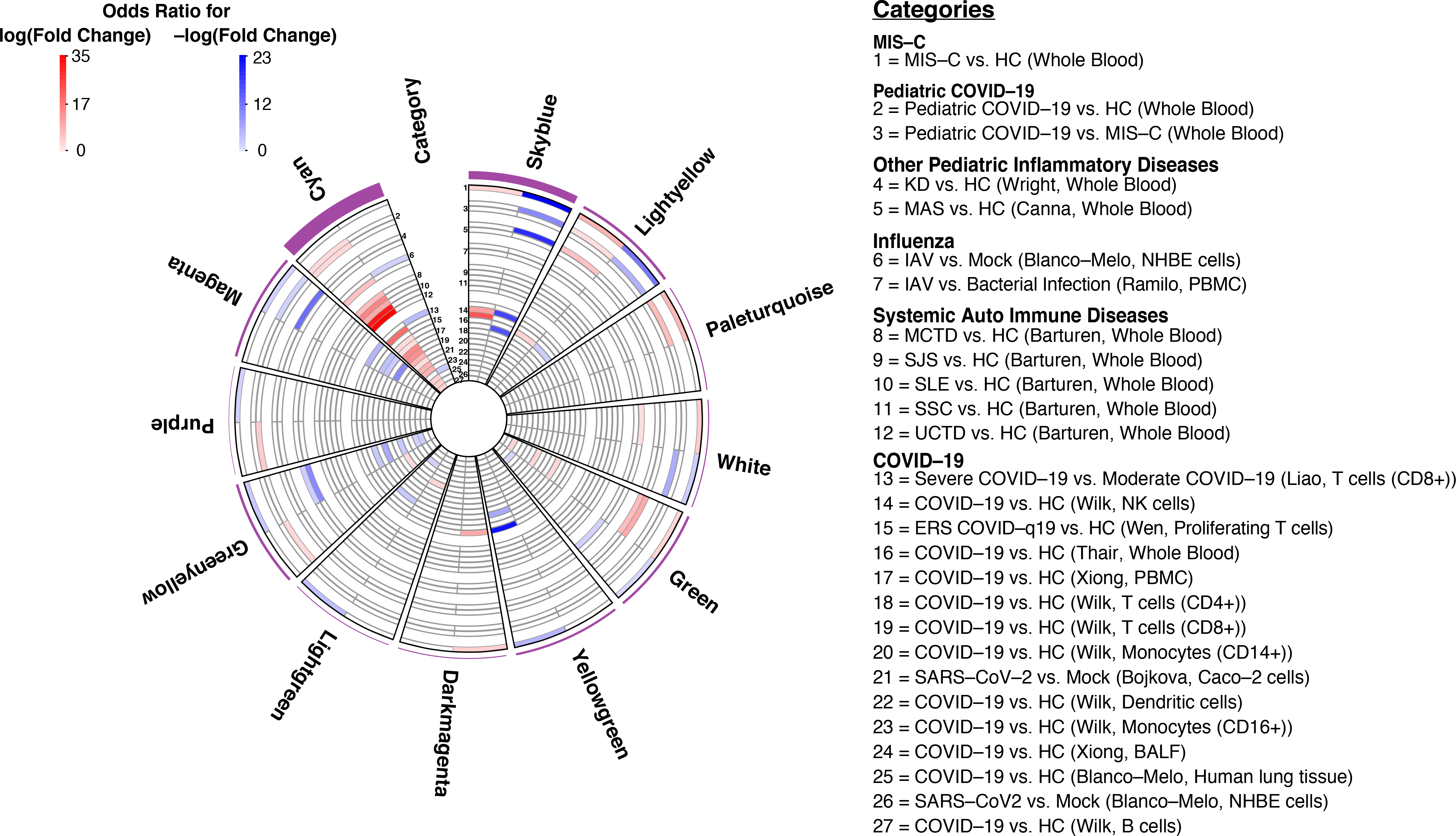
Disease enrichments in MIS-C modules and cyan. Slices are modules and each circular row represents the corresponding disease signature defined in the legend on the right. The purple outer rim of the plot represents the sum of the OR for all enrichments in that slice. Each slice is divided into two components that show the OR for enrichment of genes with +/-log(fold change) in the corresponding disease signature (red and blue respectively as defined in legend). Numbers in the category slice map the circular rows to disease signatures. OR are shown if disease signature enrichment adjusted p-value < 0.05. Diseases were grouped in biologically meaningful clusters as defined in the legend. Parenthesis in the legend refers to the source of the signature and the tissue or cell-type assessed. References for disease signatures are defined in the ***Supplementary Note*** and all enrichments can be found in ***Data S2***.

It has been proposed that MIS-C is a systemic autoimmune disease(Cavounidis et al., 2020; Consiglio et al.; Gruber et al., 2020). To address this possibility, we next projected onto the co-expression network DE signatures from a study of 918 cases and 263 healthy controls that spanned 7 classic autoimmune diseases (undifferentiated connective tissue disease, UCTD; systemic lupus erythematosus, SLE; rheumatoid arthritis, RA; mixed connective tissue disease, MCTD; systemic sclerosis, SSC; Sjögren’s syndrome, SJS; primary antiphospholipid syndrome, PAPS) (see *Methods)*. Comparing the 7 autoimmune diseases with one another, we found six modules enriched for at least two signatures, showing evidence of shared gene expression patterns across autoimmune diseases *(Data S2-4)*. Cyan showed the most ubiquitous enrichment for upregulated DEG signatures *(Figure 4;* SLE OR=10.37, FDR=1.08E-25; SJS OR=15.9, FDR=3.03E-34; SSC OR=31.4, FDR=2.40E-15; UCTD OR=34.7, FDR=1.54E-19; MCTD OR=7.29, FDR=1.46E-27), followed by darkorange (SLE OR=15.4 FDR=3.23E-22; SJS OR=11.5, FDR=1.55E-11; and MCTD OR=19.1, FDR=1.61E-48), sienna3 (SLE OR=12.3, FDR=5.53E-06; and MCTD OR=9.05, FDR=1.20E-06), and tan (SLE OR=3.93, FDR=0.0003; MCTD OR=3.60, FDR=7.26E-07). Darkolivegreen was enriched for multiple downregulated DEG signatures (SLE OR=15.8, FDR=7.77E-08; SSC OR=31.9, FDR=0.009), as was pink (SLE 3.344, FDR=0.05; MCTD OR=2.89, FDR=0.01). Comparing the autoimmune diseases with MIS-C, we found that none of the 11 modules enriched for MIS-C were enriched for autoimmune disease signatures *(Figure 4, Data S2-4)*.

### Shared gene co-expression between MIS-C and COVID-19

The timing of presentation of MIS-C and its requirement for testing positive for SARS-CoV-2 antibodies are strong, though indirect, evidence that SARS-CoV-2 infection may be a necessary but not sufficient risk factor for MIS-C. However, the molecular relationship between these two remains to be characterized. We leveraged the composition of our cohort to assess the relationship between pediatric COVID-19 and MIS-C. The pediatric COVID-19 signature was enriched in 13 modules, 5 for upregulated DEGs (cyan OR=3.24, FDR=4.40E-14; blue OR=1.99, FDR=1.60E-13; darkred OR=10.5, FDR=7.82E-51; paleturquoise OR=7.61, FDR=1.21E-17; sienna3 OR=7.07, FDR=4.18E-09), 6 for downregulated DEGs (darkolivegreen OR=5.74, FDR=2.09E-10; magenta OR=2.19, FDR=2.40E-05; orange OR=2.47, FDR=0.03; pink OR=1.69, FDR=0.05; skyblue OR=9.62, FDR=1.01E-27; turquoise OR=1.69, FDR=8.05E-07) and 2 for DEGs in both directions (black OR-Up=7.95, FDR-Up=4.07E-150, OR-Down=4.24, FDR-Down=3.17E-61; lightyellow OR-Up=2.22, FDR-Up=0.03, OR-Down=5.59, FDR-Down=3.76E-23) *(Figure 4*). Of these, 4 were also enriched for MIS-C signatures (lightyellow OR=8.86, FDR=2.83E-33; skyblue OR=3.86, FDR=0.0007; paleturquoise OR=8.31, FDR=4.12E-15; magenta OR=2.08, FDR=0.0003). To assess whether this overlap between MIS-C and pediatric COVID-19 was specific to SARS-CoV-2 infection or shared with other viral signatures, we projected influenza A virus (IAV) and Ebola virus disease (EVD) signatures onto the network (see *Methods)*. While a subset of the MIS-C modules was enriched for IAV, we did not find any enrichment for these signatures in the modules shared between MIS-C and pediatric COVID-19 *(Figure 4, Data S2-4)*.

To further corroborate the molecular link between MIS-C and SARS-CoV-2 infection, we sought to replicate and extend the module associations observed in our data to a comprehensive set of over 20 published COVID-19 DEG signatures (see *Methods, Figure 1E)*. Seventeen modules were enriched for at least 1 COVID-19 signature, with 4 modules enriched for upregulated genes, 7 modules enriched for downregulated genes, and 6 modules enriched for both *(Data S2-4)*. Seven of these were also enriched in the same direction for the MIS-C signature, and 9 of the 13 modules enriched for pediatric COVID-19 DEGs were enriched in the same direction in at least one of these published COVID-19 signatures. The cyan module stood out in this analysis, as it was found enriched for 13 of the 22 upregulated published COVID-19 signatures tested as well as the upregulated pediatric COVID-19 signature from our cohort *(Figure 4*). This module was also enriched for upregulated IAV and systemic autoimmune disease signatures, but not MIS-C *(Figure 4, Data S2-3)*. It was annotated to the GO terms ‘response to virus’ (OR=5.22, FDR=2.38E-12) and ‘response to type I interferon (OR=10.1, FDR=5.13E-12)’ and both myeloid and lymphoid cell signatures, most notably interferon-stimulated CD4+ T cells (maximum Fisher’s exact test OR=47.31, adjusted p-value=1.24 x 10-39), as well as mononuclear phagocytes (maximum Fisher’s exact test OR=6.61, adjusted p-value=4.08 x 10-11) *(Data S2-5)*.

### Skyblue implicates T-cell exhaustion in MIS-C

To further dissect the molecular traits associated to MIS-C, we focused our analyses on the co-expression network module skyblue, as it had the largest OR for enrichment of the MIS-C signature *(Figure 1F, Figure 4, Data S2-4)*. As noted above, this module was annotated to CD8^+^ T-cells and NK cells and was enriched for COVID-19 and KD signatures *(Figure 4*).

To gain a deeper understanding of how this module confers risk for MIS-C following SARS-CoV-2 infection, we leveraged cell type specific signatures associated to COVID-19 and early recovery stage following COVID-19. Although being more enriched for downregulated genes in both COVID-19 and MIS-C, the opposite effect in the skyblue module was observed for proliferating T-cells in early recovery stage, with a strong enrichment for upregulated DEGs (Fisher’s exact test OR=22.81, adjusted p-value=0.02). This suggested MIS-C as having a T-cell specific inappropriate recovery to SARS-CoV-2 infection *(Figure 4)*. To assess whether the enrichment of skyblue for early recovery stage COVID-19 DEGs was specific to CD8^+^ T-cells and NK cells (that were annotated to skyblue), we projected their signatures onto the upregulated genes in early recovery stage COVID-19 that were in skyblue. We found a significant overlap for nearly all of the CD8^+^ T-cell and NK cell signatures *(Data S2-8)*. We performed the same enrichment for CD8^+^ T-cells and NK cells onto up- and downregulated MIS-C DEGs in skyblue to definitively establish which set of DEGs was annotated to these cell types (see *Methods)*. All of the CD8^+^ T- and NK cell signatures were enriched for the subset of skyblue genes downregulated in MIS-C, and none for the upregulated subset *(Data S2-8)*, confirming that the same CD8^+^ T- and NK cell signatures upregulated in early recovery stage COVID-19 are downregulated in MIS-C. Since CD8^+^ T-cells and NK cells signatures have genes in common, we wanted to determine if one or both of these cell types were responsible for the signal observed in skyblue. To assess this, we projected onto skyblue genes present in either both CD8^+^ T-cells and NK cells, in CD8^+^ T-cells only, or genes in NK cells only. The enrichment was strongest for the genes in both cell types (OR = 43.70, p-value = 3.52E-32), but remained significant for both of the cell-specific sets (NK cells OR = 6.53, p-value = 1.66E-4; CD8^+^ T-cells OR = 3.62, p-value = 8.06E-3), suggesting both cell types independently contribute to MIS-C through skyblue *(Figure S5)*.

Next, we wanted to further annotate genes in this module to specific CD8^+^ T-cell phenotypes. During a viral infection, upon recognition of their cognate antigen, naive CD8^+^ T-cells proliferate, gain effector functions, eliminate infected cells, and either perish themselves through apoptosis or persist as memory cells that respond rapidly to future encounters with the same pathogen(Kalia and Sarkar, 2018; Wherry et al., 2007). With chronic viral infection, effector cells curtail their cytotoxic activity likely to avoid harming normal host cells, resulting in an “exhausted” functional state(Blank et al., 2019). Exhausted CD8^+^ T-cells (Tex) have been further categorized using transcriptional, epigenetic, and functional assays into four subsets along the differentiation path of these cells: two progenitor states (Tex^prog1^ and Tex^prog2^), an intermediate state where certain effector-like functions are re-acquired (Tex^int^) and a terminally exhausted state (Tex^term^)(Beltra et al., 2020). Transcriptional signatures were obtained from the literature for Tex compared to effector and memory CD8^+^ T-cells(Wherry et al., 2007) and for the four Tex subsets compared to one another(Beltra et al., 2020)(see *Methods)*. Enrichment in skyblue was observed for DEGs upregulated in Tex compared to both memory (Fisher’s exact test 0R=10.23, adjusted p-value=7.11 x 10-4; *Figure 5A, Data S2-8* and effector subtypes (Fisher’s exact test 0R=6.96, adjusted p-value=0.03; *Figure 5A, Data S2-8)*. To test if this observation was specific to skyblue or characteristic of all MIS-C CD8^+^ T-cell modules, we performed the same enrichment tests in the magenta module (also downregulated in MIS-C and annotated to CD8^+^ T-cells) and found the opposite effect, enrichment of genes upregulated in effector (Fisher’s exact test 0R=2.90, p-value=0.01) and memory (Fisher’s exact test 0R=3.18, p-value=0.006) CD8^+^ T-cells compared to Tex *(Figure S5)*. Between the Tex subsets, we found significant enrichment of the signatures for Tex^int^ and Tex^term^ compared to the Tex^prog1^ and Tex^prog2^ *(Figure 5A)*, indicative of more mature Tex gene expression in skyblue. Taken together, these observations pinpoint the downregulation of mature exhausted CD8^+^ T-cells in the molecular pathology of MIS-C.

**Figure 5:**
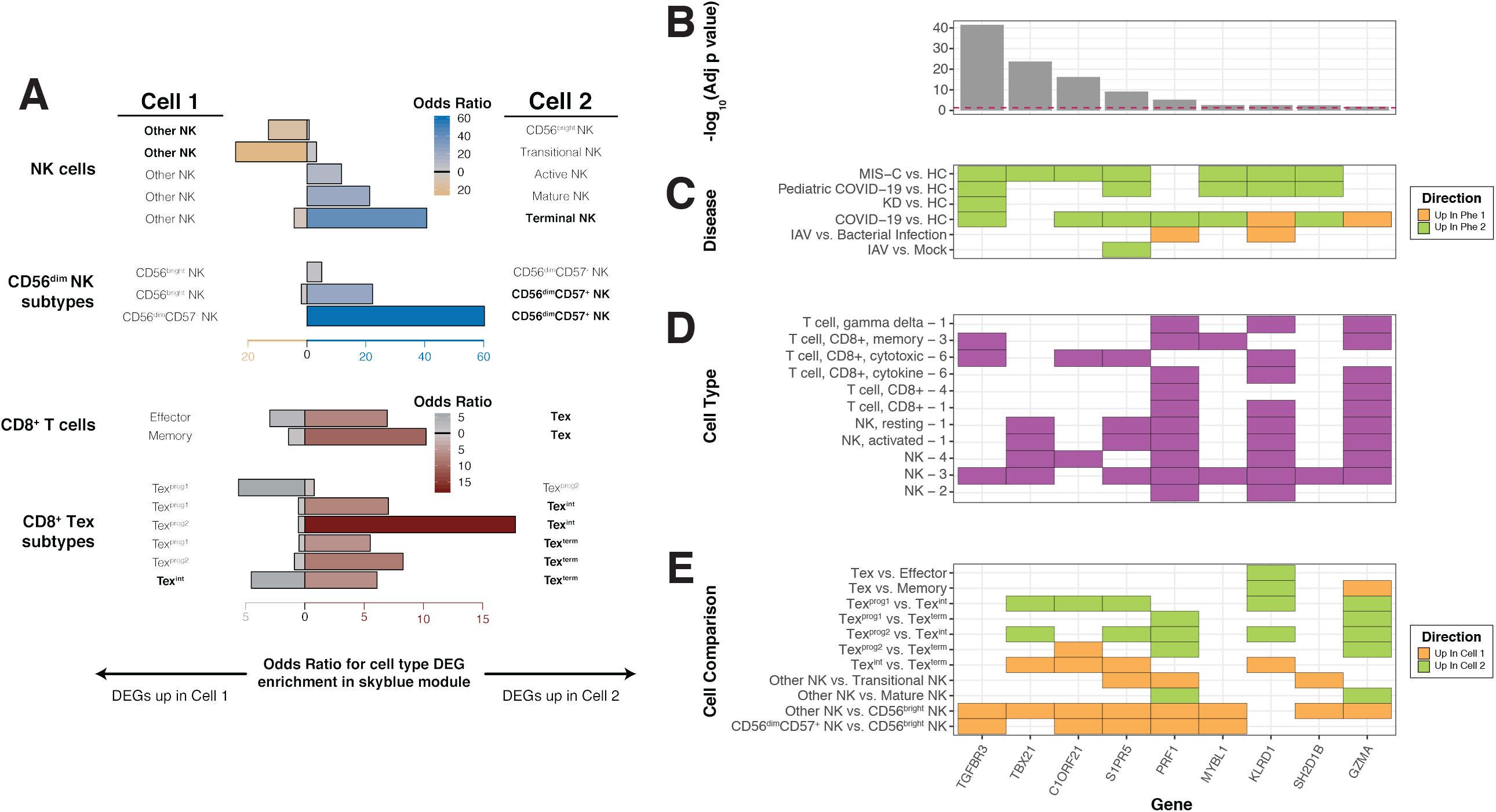
Further dissection of skyblue implicates cytotoxic lymphocyte subtypes in MIS-C. A: Defining NK cell and CD8^+^ T-cell subtypes in skyblue. The x-axis represents the OR for the enrichment of skyblue for specific cytotoxic cell subtypes and the y-axis the signatures used. Cell one and cell two refer to the cell subtypes being compared to generate the DE signatures projected onto skyblue and the direction of the bar is shown towards the upregulated cell type in the comparison. The color and height of bars represent the OR of the enrichment test as defined by the x-axis and the legends. For NK cells, we used signatures from Yang, et al, Nature Communications, 2019 (labeled NK cells), and from Collins et al, Cell, 2019 (labeled CD56^dim^ NK subtypes). For CD8^+^ T-cells, we used signatures from Wherry, et al, Immunity, 2007 (labeled CD8^+^ T-cells), and from Beltra et al, Immunity, 2020 (labeled CD8^+^ Tex subtypes). Detailed descriptions of the subtypes can be found in the ***Supplementary Note***. B,C,D,E: Skyblue key drivers measures. These panels share their x-axis that represents the key driver genes. B: Key driver analysis results. The y-axis of this panel is the -log_10_(adjusted P-value) of key driver analysis. The red dashed line is the significance threshold at - log_10_(0.05). C: Related disease differential expression. In this panel, the y-axis is the disease name corresponding to Figure 4. The color represents the direction in the comparison as defined in the legend. D: Cell-type specific expression of key driver genes. The y-axis is the cell type signatures from the following references: 1= Newman et al, Nature Methods, 2015; 2= Park, et al, Science, 2020; 3=Wilk, et al, Nature Medicine, 2020; 4= Lial, et al, Nature Medicine, 2020; and 6= Szabo, Nature Communications, 2019. E: CD8^+^ T-cell and NK cell subtypes specific signatures. The y-axis is the cell-type name corresponding to Figure 5A and the color represents the direction in the comparison as defined in the legend.

**Figure 6:** Causal information flow between MISC modules and cyan. Genes are shown if they were key drivers of a target module as well as upstream of a key driver gene in the target module for any of the MIS-C modules or cyan. Circles represent key driver genes and rectangles target modules, and genes and modules are colored by module names. Causal relationships between genes and modules are depicted by an arrow that is colored by the parent module name.

We next assessed whether specific subtypes or differentiation states of NK cells were representative of the genes in skyblue. There are 2 types of NK cells: CD56^bright^ and CD56^dim^. CD56^bright^ NK cells are less common, less cytotoxic, and more active at secreting cytokines to trigger immune responses(Michel et al., 2016). CD56^dim^, in contrast, make up 90% of NK cells in blood and are highly cytotoxic(Michel et al., 2016). To establish which class of NK cells contributes to MIS-C, we projected subtype-specific NK cell signatures from 3 independent datasets onto skyblue (see *Methods)*. In all references tested, skyblue was enriched for genes up in CD56^dim^ and down in CD56^bright^, thereby showing the NK cells contributing to MIS-C pathology are of the CD56^dim^ subtype *(Figure 5, Data S2-8)*. CD56^dim^ NK cells can be further split into 2 subtypes: CD57+ and CD57^-^. CD56^dim^CD57+ NK cells comprise 30-60% of CD56^dim^ NK cells(Lopez-Vergès et al., 2010) and have adaptive-like features (i.e., memory in response to particular viruses) compared to CD56^dim^CD57^-^ NK cells(Nielsen et al., 2013). One of the datasets we tested provided signatures for CD56^dim^CD57+ NK cells compared to CD56^dim^CD57^-^ NK cells and compared them to CD56^bright^ NK cells(Collins et al., 2019). Projection of these comparisons onto skyblue showed that the enrichment for CD56^dim^ NK cells over CD56^bright^ NK cells was explained by CD56^dim^CD57+ NK cells (Fisher’s exact test 0R=22.27, adjusted p-value=1.12 x 10-16;) and that skyblue was significantly enriched for the CD56^dim^CD57+ NK cell signature compared to CD56^dim^CD57^-^ (0R=60.17; adjusted p-value=7.61 x 10-12, *Figure 5A, Data S2-8)*. Together, these results determine that genes upregulated in cytotoxic CD56^dim^CD57^+^ NK cells are enriched in skyblue and are therefore downregulated in MIS-C.

### MIS-C modules projected onto Bayesian networks

Whereas co-expression networks primarily capture linear relationships between genes, they do not account for more complex, nonlinear and statistically inferred causal regulatory relationships. Bayesian networks (BNs) are graphical models useful for the latter and have proven a valuable framework for capturing the flow of information within high-dimensional molecular data. We used BNs to better understand how the modules implicated in MIS-C are regulated *(Figure 1E, 1F)*. The rarity of MIS-C did not allow sufficient samples to construct a causal network for this disease directly. Instead, we identified a recently constructed BN from a large IBD cohort comprised of 209 healthy controls, 389 ulcerative colitis patients, and 432 Crohn’s disease patients from whom blood was collected and RNA sequenced(Suárez-Fariñas et al., 2020). We examined the 11 MIS-C modules and cyan by projecting them onto the BN, identifying all genes within one path length of these module genes and the largest connected subnetwork from these projections as the representation of the module in the BN. All of the MIS-C modules were observed to be well conserved in the BN. For example, the subnetwork identified for the skyblue module was more than 29-fold enriched for genes in the skyblue module (p-value=1.5e-47), demonstrating strong conservation between the topology of the skyblue module and the blood-derived BN.

We next performed key driver analysis on each of the MIS-C module subnetworks to identify those genes predicted by the BN to regulate the module *(Data S5-1)*. Key driver analysis was performed using the Key Driver Analysis R package(Zhang and Zhu, 2013) in which a key driver is defined as a gene that is significantly upstream and connected to a set of genes of interest (i.e., a module). For the skyblue module we identified 9 key driver genes *(TGFBR3, TBX21, C1ORF21, S1PR5, PRF1, MYBL1, KLRD1, SH2D1B*, and *GZMA)* where changes in the expression of these genes are predicted to alter the regulatory state of the module. We annotated these key drivers, showing the strength of the enrichment in the key driver analysis *(Figure 5B)*, the DE direction within disease states *(Figure 5C)*, the membership to a cell type specific signature *(Figure 5D)*, and the DE direction within T-cell and NK cell subsets *(Figure 5E)*. We observed that these key drivers were generally downregulated in MIS-C and other diseases, often markers of CD8^+^ T-cell and NK cell subtypes, and frequently expressed in later stages of CD56^dim^CD57+ NK and Tex differentiation. Finally, while the most significant key driver in terms of its regulatory impact on the skyblue module was *TGFBR3*, the second highest impact key driver was *TBX21*, where changes in the expression of this gene are predicted to impact 114 genes in the BN, 24 of a which reside in the skyblue subnetwork, a 25.2-fold enrichment over what would be expected by chance (adjusted p-value = 2.3e-28). *TBX21* was recently characterized as a biological switch that facilitates the transition of exhausted CD8^+^ T cells in a progenitor state to an intermediate state on the way to terminal exhaustion(Beltra et al., 2020), consistent with our observation of the skyblue module implicating CD8^+^ T-cell exhaustion as a hallmark of MIS-C.

## Discussion

In this study, we present what is to our knowledge the first genome-wide investigation of the molecular etiology of MIS-C. We quantified gene expression changes in MIS-C and pediatric COVID-19 cases compared to HCs in 30 whole blood samples from 19 individuals. Cell type deconvolution found reduced proportions of circulating CD8^+^ T-cells in MIS-C cases, and DE between MIS-C cases and HCs defined “the MIS-C signature” as genes with nominally significant p-values. Upregulated genes in this signature were annotated to both myeloid and lymphoid immune functions, while downregulated genes were annotated to pathways related to the regulation of gene expression. We built a co-expression network from the 30 samples in our cohort to further dissect the MIS-C signature, harnessing its structure to probe the predominant hypotheses regarding MIS-C pathogenesis, including its overlap with KD, its molecular link to SARS-CoV-2 infection, and its similarity to classic autoimmune diseases. By projecting the MIS-C signature onto this network we identified a set of 11 modules enriched for DEGs. These modules were also enriched for KD and COVID-19 signatures, but not for autoimmune disease signatures. The skyblue module showed the strongest enrichment for downregulated MIS-C DEGs and was specifically enriched for signatures of exhausted CD8^+^ T-cells and CD56^dim^CD57+ NK cells, pinpointing their downregulation as important in MIS-C. Finally, probabilistic causal network modeling prioritized *TBX21* and eight other genes as key drivers of MIS-C pathogenesis.

This study has several limitations. Due to the rarity of MIS-C and the urgency of the situation, we were only able to capture transcriptomic information for eight MIS-C cases. With fewer than 600 cases reported nationally at the time of writing(Godfred-Cato, 2020), the current cohort represents a non-trivial proportion (1.4%) of the total population afflicted to date in the United States. We did not detect genome-wide significant DEGs between MIS-C and HCs at this sample size, however, using L_1_ statistics, we estimated that ~15% of genes in the analysis are true DEGs. These observations suggest we were sufficiently powered to detect a transcriptome-wide MIS-C signature despite being underpowered to detect individual DEGs, which was further corroborated by the molecular overlap observed between MIS-C and KD. Another limitation is MIS-C cases and HCs were not matched for age or ethnicity. HCs were individuals working on-site who were asymptomatic for COVID-19. This was the most optimal study design that could be achieved given the study occurred in New York City at the local height of the COVID-19 pandemic. We mitigated the age mismatch using gene expression changes known to be associated with age, developing an approach that allowed us to account for the age contribution to gene expression without removing the disease signal. It was not possible to perform a similar correction for the ethnicity mismatch due to the lack of published ethnicity-specific blood expression signatures. Finally, our pediatric COVID-19 comparator group had immunological comorbidities (e.g., B-cell ALL, IBD) that likely impacted their transcriptomic profiles. That said, we found that the COVID-19 signature derived from these individuals when compared to HCs was largely consistent with published COVID-19 signatures from adult cases and controls without such comorbidities. Altogether, utilizing a large amount of data in a hypothesis-driven fashion allowed us to partially overcome the main limitations of our study and gain meaningful insights despite suboptimal design.

One of the most pressing questions regarding MIS-C is its relationship to KD. The presence of conjunctival injection, oral erythema, rash, and other symptoms often seen in KD initially prompted MIS-C to be termed the “Kawasaki-like disease”(Verdoni et al., 2020). However, the two conditions notably differ with respect to age of presentation and ethnicity of the affected population. The incidence of KD peaks at 10 months of age(Rowley, 2020), whereas 70% of MIS-C cases occur in children over 6 years old(Dufort et al., 2020). Additionally, individuals of Asian descent are consistently found to have the highest incidence of KD(Uehara and Belay, 2012), while early epidemiological reports of MIS-C have found incidence highest amongst individuals self-reporting as Black or Hispanic(Dufort et al., 2020; Riphagen et al., 2020; Toubiana et al., 2020). Our analyses provide molecular support for the clinical intuition that MIS-C shares similarities with KD. Most notably, the skyblue module was critically downregulated in both conditions. This suggests a common molecular foundation between the two that was not observed between MIS-C and other pediatric multisystem immune disease signatures evaluated. Our results also support the notion that, while similar, MIS-C and KD are not necessarily the same disease. The paleturquoise module, for instance, showed a strong enrichment for upregulated genes in MIS-C, platelet markers, and coagulation pathways. The lack of enrichment for the KD signature in paleturquoise leads to the hypothesis that KD coagulopathies(Burns et al., 1984) may occur via different molecular mechanisms than MIS-C coagulopathies. Overall, the molecular comparison between MIS-C and KD emphasized similarities and differences that may account for their being clinically alike yet distinct.

In addition to being linked to KD, two recent reports (including one analyzing the cohort presented here)(Cavounidis et al., 2020; Consiglio et al.; Gruber et al., 2020), put forth evidence that MIS-C shares molecular pathophysiology with classic autoimmune diseases. We performed a transcriptome-wide assessment of this hypothesis, evaluating whether the architecture of co-expression was shared between classic autoimmune diseases and MIS-C. Strikingly, we found that no co-expression modules were enriched for both MIS-C and autoimmune disease signatures using the most comprehensive RNA sequencing study of autoimmune diseases performed to date(Barturen et al., 2020), suggesting that at the level of gene expression in the blood there is little overlap. This does not rule out a role for autoantibodies in disease pathogenesis, but rather may be indicative of an autoimmune mechanism unique to MIS-C.

A key unknown aspect of MIS-C pertains to its relationship to SARS-CoV-2 infection, where a causal relationship is suggested by the timing of MIS-C case clusters following local surges in COVID-19 and the presence of SARS-CoV-2 antibodies in the sera of MIS-C cases(Verdoni et al., 2020). We have identified a link between SARS-CoV-2 infection and MIS-C at the level of gene expression, with several modules enriched for MIS-C and both pediatric and adult COVID-19 DE signatures. This does not establish causality, but provides evidence of a direct molecular link between MIS-C and both the acute and convalescent stages of SARS-CoV-2 infection.

Though our work was primarily focused on MIS-C, we also identified the cyan module as central to the molecular pathology of COVID-19. This module was enriched for both our pediatric COVID-19 upregulated DEGs as well as 13 of the 22 published COVID-19 upregulated signatures included in our analyses. Both pathway and cell type annotations linked this module to the interferon response, substantiating the hypothesis that imbalanced interferon response is a hallmark of COVID-19(Acharya et al., 2020). This finding could have clinical implications. Due to the observation of a blunted interferon response during the incubation phase of SARS-CoV-2 infection(Blanco-Melo et al., 2020), experimental prophylactic treatment with interferons has already been used in at-risk human subjects(Meng et al.). Our findings do not preclude the use of interferons in this manner, where priming the immune system to a pre-existing antiviral state could prevent the infection. However, individuals showing signs and symptoms of COVID-19 are likely past the point where such treatments would be beneficial, as the infection has already taken hold. Our findings suggest that the interferon response is upregulated in these individuals as their immune system attempts to fight off the virus, in which case this treatment would exacerbate their condition. Indeed, this has been found in animal models of other (3-coronaviruses(Channappanavar et al., 2019). A dual approach to experimental therapeutic development should therefore be considered, with interferon agonism to prevent SARS-CoV-2 infection and interferon antagonism to treat COVID-19.

The skyblue module had the strongest enrichment for MIS-C DEGs. This module may capture molecular processes that are necessary for effective elimination of SARS-CoV-2, as suggested by its being enriched for downregulated DEGs of the acute COVID-19 presentation and upregulated DEGs of the early recovery stage of COVID-19. Six of the eight MIS-C cases in our dataset tested negative for SARS-CoV-2 infection on presentation, indicating successful clearance of the virus. Yet, in MIS-C, skyblue was most strongly enriched for downregulated DEGs, the same direction seen in the acute COVID-19 presentation and opposite to what is observed in the early recovery stage, perhaps reflecting an improper recovery from SARS-CoV-2 infection leads to MIS-C. Specifically, the cell type signature and pathway enrichment analyses of skyblue implicated the downregulation of exhausted CD8^+^ T-cells and CD56^dim^CD57^+^ NK cells in MIS-C. While these cell types share transcriptional signatures, our results suggest both independently contribute to disease. The predominant cytotoxic cells of the immune system, CD8^+^ T-cells and NK cells are equipped to kill infected or otherwise compromised cells and operate in synchrony, interacting directly and indirectly to co-regulate one another(Uzhachenko and Shanker, 2019). NK cell depletion is known to prevent CD8^+^ T-cell exhaustion(Cook and Whitmire, 2013; Waggoner et al., 2011), and the absence of NK-dependent clonal expansion of CD8^+^ T-cell exhaustion after viral infection can lead to severe and even fatal T-cell immunopathology(Cook and Whitmire, 2013; Waggoner et al., 2011). These latter observations are consistent with other reports of inflammatory disease symptoms improving with CD8^+^ T-cell exhaustion(McKinney et al., 2015).

We used an available BN(Suárez-Fariñas et al., 2020) to determine the regulatory structure of skyblue, organizing whole blood gene expression from individuals with IBD into a probabilistic causal network wherein 9 key drivers of skyblue were identified *(TGFBR3, TBX21, C1ORF21, S1PR5, PRF1, MYBL1, KLRD1, SH2D1B*, and *GZMA)*. All of these genes have known associations with CD8^+^ T-cell and NK cell functions(Drouillard et al., 2018; Foltz et al., 2018; He et al., 2016; Jenne et al., 2009; Leavy, 2012; Naluyima et al., 2019; Roncagalli et al., 2005; Tinoco et al., 2009; Wang et al., 2019; Yeo et al., 2018) and a subset have been associated with illnesses that share clinical presentations with MIS-C. These include *PRF1* in hemophagocytic lymphohistiocytosis(Kim et al., 2014; Lee and Molleran Lee, 2004), *MYBL1* in chronic active Epstein-Barr virus infection(Zhong et al., 2017), *KLRD1* in influenza symptom severity(Bongen et al., 2018) and mousepox(Fang et al., 2011), and *SH2D1B* in SLAM family receptors in immune disorders(Cannons et al., 2011). Of the 9 key drivers, *TBX21* is of particular interest due to the link our other analyses established between Tex cells and MIS-C. The enrichment of skyblue for downregulated MIS-C DEGs and for Tex^int^ and Tex^term^ signatures suggests CD8^+^ T-cells fail to reach mature exhausted states in this disease. *TBX21* codes for the transcription factor T-bet, which in a recent report(Beltra et al., 2020) was found to be the primary coordinator of the conversion of Tex^prog2^ to Tex^int^. This conversion was also the Tex subset comparison in our analyses that showed the strongest signal in skyblue *(Figure 5)*, with enrichment seen for genes upregulated in Tex^int^ compared to Tex^prog2^. Our identification of *TBX21* as downregulated in MIS-C compared to HCs and a key driver of skyblue, combined with the central role *TBX21* is known to play in Tex maturation and the skyblue enrichment for mature Tex signatures, are compelling clues that this gene plays a central role in the molecular etiology of MIS-C.

In summary, with this report we aimed to characterize the molecular pathogenesis of MIS-C and found dysregulation of cytotoxic lymphocytes as a hallmark of the condition. Its downregulation in MIS-C supports cytotoxic T-cell exhaustion as a normal physiological condition, not simply a pathological state that occurs in cancer and chronic infection(Blank et al., 2019). The link to CD8^+^ T-cell exhaustion was not just seen in MIS-C but in KD as well, a disease whose etiology remains elusive. Just as therapeutic approaches reversing exhaustion are actively being explored to treat cancer(Marro et al., 2019), approaches that induce exhaustion may be effective for diseases such as MIS-C where its absence is pathological. *TBX21* and the other key drivers of the skyblue module are promising therapeutic targets in this regard. Though our study is underpowered to define SARS-CoV-2 infection as causal to the molecular processes in MIS-C, we show that they share molecular features, suggestive of a causal chain. Understanding the link between SARS-CoV-2 and MIS-C may have ramifications beyond pediatric cohorts. A subset of adults who recover from COVID-19 report symptoms such as fatigue and dyspnea that linger for months(Carfì et al., 2020) and we hypothesize that cytotoxic lymphocyte dysregulation as observed in MIS-C may be a contributing factor in such cases. As independent cohorts are assembled, replication and validation of our primary findings in blood and other tissues will be paramount.

## Data Availability

Data will be made available upon request following publication in a peer-reviewed journal

## STAR Methods

### RNA Extraction

Peripheral blood was collected from study participants into Tempus RNA Blood Tubes (ThermoFisher, #4451893). As soon as possible after blood collection, tubes were stored at -80°C until RNA extraction. The MagMax protocol for Stabilized Blood Tubes RNA Isolation Kit (Ambion, Life Technologies) was used to extract total RNA from 3 mL of collected peripheral blood, following the manufacturer’s instructions. Briefly, frozen blood tubes were thawed to 4°C prior to RNA extraction. In a biosafety level 2+ (BSL2+) cabinet using proper personal protective equipment (PPE), the blood was first diluted with 1X PBS, then pelleted and washed at 4°C. The washed and re-pelleted RNA was then dissolved in Resuspension Solution followed by a dual proteinase and DNase treatment. Next, the RNA was purified through a series of magnetic bead-based washes and eluted in 80 |jl of Elution Buffer as described in the user manual, prior to downstream quantification and library preparation steps. In addition, these modifications to the protocol were followed during extraction: 1) all processing was performed inside a BSL2+ cabinet until resuspension solution was added to the samples, 2) samples were kept on ice throughout the protocol, 3) wash 1 buffer, wash 2 buffer, and isopropanol were kept on ice and used cold, 4) samples were shaken on a vortex adaptor at a setting of 4.

### RNA-seq library preparation and sequencing

Total RNA was examined for quantity and quality using the Fragment Analyzer (Agilent) and Quant-It RNA (ThermoFisher) systems. RNA samples with sufficient material (>50 ng) were passed to whole-transcriptome library preparation using the TruSeq Stranded Total RNA Library Prep Gold (Illumina) following the manufacturer’s instructions. The kit is specifically designed to remove highly abundant ribosomal, globin and mitochondrial RNA (r-globin- and mt-RNA) from whole blood-derived total RNA in order to enrich for both protein-coding and non-coding RNAs, including microRNAs. Briefly, total RNA inputs were normalized to 100ng/10^L (if 100ng were not available, input amounts were utilized as low as 50ng) going into preparation. Total RNA was first depleted of highly abundant rRNA, mtRNA and globinRNA species, prior to enzymatic fragmentation and cDNA generation. The 3’ ends of cDNA were then adenylated prior to ligation with adapters utilizing unique dual indices (96 UDIs) to barcode samples to allow for efficient pooling and high throughput sequencing. Libraries were enriched with PCR, with all samples undergoing 15 cycles of amplification prior to purification and pooling for sequencing. Completed libraries were quantified using Quant-iT reagents and equimolar pools were generated and sequenced on the NovaSeq 6000 using Sprime flow cells with 100 base pair paired-end reads, generating a mean of 50 million paired-end reads per sample.

### RNA-seq data processing, quality control and normalization

Once sequencing data completed collection on the instrument, and base calls were converted into raw reads, these raw reads were filtered after quality assessment. The quality-filtered raw data was then converted into FASTQ files using bcl2fastq Conversion tool (Illumina). RNA-seq reads were aligned to the GRCh38 primary assembly with Gencode release annotation by STAR (v2.7.3a)(Dobin et al., 2013) using per-sample 2-pass mapping (--twopassMode

Basic) and chimeric alignment options (--chimOutType Junctions SeparateSAMold -chimSegmentMin 15 - chimJunctionOverhangMin 15). RNA-seq QC metrics were calculated by fastqc (v0.11.8) and Picard Tools (v2.22.3)(Broad Institute). Quantification was done at the gene-level with STAR (--quantMode GeneCounts), at the transcript level with kallisto (v0.46.1), and with antisense specificity using featureCounts(Liao et al., 2014) (Subread R package v1.6.3 and strandness option -s 2) for various counting criteria, including gene-level grouping (-t exon -g gene_id), gene-level grouping / primary alignments only / count all overlapped features (-t exon -g gene_id -primary -O), transcript-level grouping (-t exon -g transcript_id), and exon level / count all overlapped features (-t exon -f -O). A customized version of MultiQC(Ewels et al., 2016) was used the compile and summarize the per-sample statistics from STAR, Picard Tools and featureCounts (i.e. gene-level counts, mtRNA counts, globinRNA counts, etc) into an interactive HTML report.

We next assessed whether there was mislabeling in our cohort. To do so, we combined two levels of information: correspondence of imputed gender from the gene expression data to the clinical data *(Figure S1A)* and identity concordance using genetic information derived from the sequencing data using NGSCheckMate(Lee et al., 2017). Using these, we were able to confirm that no samples were mislabeled. After removing lowly expressed genes (keeping genes with counts per million > 1 in 10% of the samples), we normalized the raw count data of the 22,302 remaining genes using voom from the limma R package(Ritchie et al., 2015). One sample failed to pass all quality controls and was removed from further analyses. Principal components analyses to explore covariate effect on gene expression variance genome-wide were done using the prcomp R function. Canonical correlation analyses were performed and plotted with the canCorPairs and plotCorrMatrix function from the variancePartition R package(Hoffman and Roussos, 2020; Hoffman and Schadt, 2016), and permutation p-values were computed with the p.perm function of the CCP R package(Menzel, 2012). The normalized expression data was adjusted for the following covariates using the fitVarPartModel and the get_predictions function of the variancePartition R package version 1.19.6(Hoffman and Roussos, 2020; Hoffman and Schadt, 2016) (Batch, median insert size, RNA Integrity Number, percent chimeras, PF HQ error rate and median CV coverage and sex). The median CV coverage is defined by Picard Tools(Broad Institute) as “the median coefficient of variation (CV) or stdev/mean for coverage values of the 1000 most highly expressed transcripts” and the PF HQ error rate is defined as “the fraction of bases that mismatch the reference in PF HQ aligned reads.” The age cross-module eigengene was also included as a covariate in the models. Batch effect was calculated at a per gene basis using technical replicates sequenced in all batches. Technical replicate measures were combined in a single gene expression vector by removing the effect of covariates described above followed up by adding back individual effects in the linear model.

### Cell-type deconvolution

Cell type deconvolution was performed with the Cibersortx software, using transcripts per million as input and following procedures recommended by the developers(Steen et al., 2020). To ensure that we capture signals that are driven by cell type compositions and not by any single reference used, we used 3 references generated independently (i.e., by different groups, with different technologies) for this analysis. The LM22 reference(Newman et al., 2019) was generated by sorting PMBCs with fluorescent activated cell sorting (FACS) and performing bulk RNA-seq. The NSCLC PMBC(Newman et al., 2019) and SCP424(Ding et al., 2019) references were generated by single-cell RNA-seq experiments from PBMCs. For SCP424, prior processing had resulted in an unacceptably large range in total UMI present in published cells, with the range spanning over three orders of magnitude (17 to 53,562 total UMI in individual cells). We therefore removed cells with less than 630 observed genes or less than 1584 total UMI. Furthermore, to account for differences in chemistry or capture efficiency across cells, we performed the simple but lossfull procedure of UMI downsampling to bring all cells to the same level of 1584 total UMI.

Validation of compositions was performed using Pearson correlation (cor.test R function) between measured complete blood counts and corresponding deconvolved cell type fraction. For each reference, p-values were adjusted for multiple testing using the false discovery rate (FDR) estimation method of Benjamini-Hochberg(Hochberg and Benjamini, 1990) for the number of validation tests in the reference. Lymphocytes compositions were computed by adding fractions of all T- and B-cell populations. Specifically, for NSCLC, we added fraction estimates for T cells CD8, T cells CD4 and B cells; for SCP424 CD4+_T_cell, Cytotoxic_T_cell and B_cell; and for LM22 B cells naive, B cells memory, Plasma cells, T cells CD8, T cells CD4 naive, T cells CD4 memory resting, T cells CD4 memory activated, T cells follicular helper, T cells gamma delta and T cells regulatory (Tregs). For SCP424, monocyte composition was computed by adding fractions of CD16+_monocyte and CD14+_monocyte.

We used the lm function in R to test for differences in cell type compositions between disease groups, while adjusting for the age cross-module eigengene and for sex. For each cell type in each reference, pairwise comparisons were performed between MIS-C, pediatric COVID-19 and HCs. For each of phenotype comparisons, p-values were adjusted for multiple testing separately for each reference using the false discovery rate (FDR) estimation method of Benjamini-Hochberg(Hochberg and Benjamini, 1990) for the number of cell types profiled in the reference.

### Differential Expression

Differential expression (DE) analyses were performed with the dream function from the variancePartition R package(Hoffman and Roussos, 2020; Hoffman and Schadt, 2016). The covariates included in the model were sequencing batch, individual, median insert size, RNA integrity number, sex, percent chimeras, PF HQ error rate, median CV coverage, and the age cross-module eigengene. A total of 3 DE signatures were generated (MIS-C vs. control, MIS-C vs. pediatric COVID-19, and pediatric COVID-19 vs. control). Multiple testing was controlled separately for each DE comparison accounting for the 22,302 genes tested using the false discovery rate (FDR) estimation method of Benjamini-Hochberg(Hochberg and Benjamini, 1990).

### Co-expression network analyses

All co-expression analyses were performed using weighted correlation network analysis (WGCNA) R package version 1.69(Langfelder and Horvath, 2008). Co-expression network modules were created using the residual gene expression of all 30 samples after accounting for biological and technical covariates described above. Standard practices were followed to construct this network, with a soft-power threshold of 8 and a minimum module size of 30 genes.

### Pathway enrichment analyses

Enrichment for GO, C7, and Hallmark pathways (N=10,192, 4872, and 50 pathways, respectively) was carried out for the upregulated and downregulated signatures from three DE analyses (MIS-C compared to HCs, pediatric COVID-19 compared to HCs, and MIS-C compared to pediatric COVID-19) and the N = 37 modules from the co-expression network, for a total of N = 43 feature sets of interest. For each pathway in each database, enrichment was tested as the overlap between the genes in the pathway and the genes in the feature set using as background the 22,302 genes expressed in the dataset. GO analyses were done using the R packages goseq, topGO and org.Hs.eg.db while C7 and Hallmark analyses were done using the R packages HTSanalyzeR(Wang et al., 2011), GSEABase, and GAGE(Luo et al., 2009). For each feature set, multiple testing correction for the number of pathways was carried out separately for each pathway database using the false discovery rate (FDR) estimation method of Benjamini-Hochberg implemented in R using the p.adjust() function.

### Module enrichment for cell types and disease expression signatures

To evaluate whether co-expression modules captured the pathology of MIS-C and other related diseases, we utilized several reference datasets from the literature, each intended to provide a different level of insight into the molecular biology at play within our network, that are described in the *Supplementary Note*. A total of 38 disease DE signatures were evaluated: 3 signatures from our data (MIS-C vs. HCs, MIS-C vs. pediatric COVID-19, pediatric COVID-19 vs. HCs) and 35 published disease signatures. The upregulated and downregulated DE signatures were projected onto the 38 (37 modules and the set of genes that did not fall into a module in our co-expression network separately), with the exception of 1 dataset (Gordon et al, which only had an upregulated disease signature for COVID-19). This totaled 75 disease DE signatures tested. Thus, study-wide, a total of N = 2,850 tests of disease signature enrichment in co-expression modules were performed.

To evaluate whether co-expression modules were representative of the activity of specific cell types, we utilized several reference datasets from the literature, each intended to provide a different level of insight into the molecular biology at play within our network, that are described in the *Supplementary Note*. A total of 116 cell types expression profiles from the literature were evaluated for enrichment in the 37 modules and the set of genes that did not fall into a module in our co-expression network separately. Only an upregulated signature was provided for all but 1 cell type, for a total of 117 cell type DE signatures tested. Therefore, study-wide, we performed 4,446 tests of enrichment for cell type signatures in co-expression network modules.

For every combination of module and DE signature (whether disease or cell type DE signature), we performed a Fisher’s exact test to evaluate enrichment of the module for the DEGs using all 22,302 genes expressed in our dataset as background, with the null hypothesis being that module and DE signature membership are independent. To account for multiple tests we used a stringent two-step process that was performed separately for the disease and cell type DE signature analyses. First, we used a Bonferroni correction to adjust the Fisher’s exact test p-value for the number of modules (N = 37 plus 1 group of genes that did not fall into any module) against which each DE signature was tested. We did so because gene-module memberships are unique and that therefore all enrichments tests are independent from one another. These adjusted p-values were further corrected for the number of DE signatures tested (N = 75 for the disease enrichments and N = 117 for the cell type enrichments) using the method of Benjamini-Hochberg to control the false discovery rate. Here, unlike for gene-module memberships, DE signatures are highly correlated since we tested multiple signatures of the same condition (i.e., COVID-19 signatures from multiple sources) or cell type (e.g., CD8^+^ T cells from multiple sources).

To delineate whether the cell types annotated to skyblue were limited to one MIS-C signature direction, we defined “MIS-C upregulated” and “MIS-C downregulated” subsets of skyblue by keeping the genes in the module upregulated or downregulated in MIS-C, respectively. We then projected the CD8^+^ T-cell and NK cell signatures that we had found enriched in skyblue earlier onto the upregulated and downregulated subsets. As these were targeted secondary analyses, we did not perform multiple test corrections.

In addition to the primary cell type enrichment analyses, we performed hypothesis-driven follow-up analyses to further delineate the subtypes of CD8^+^ and NK cells enriched in the skyblue module. For these analyses, since they were targeted hypotheses for a single module, we only adjusted p-values for the number of DE signatures tested (N = 43) using the method of Benjamini and Hochberg.

### Bayesian Network analysis

In the context of a large independent IBD study, whole blood RNA-seq data was obtained via a protocol approved by the Icahn School of Medicine at Mount Sinai Institutional Review Board. This data was representative of a cross sectional cohort who were enrolled in the Mount Sinai Crohn’s and Colitis Registry (MSCCR) between December 2013 and September 2016. This data and the analyses described below will be presented in an independent publication (in review). Informed consent was obtained from all participants recruited into this independent study. The MSCCR is composed of patients with Crohn’s disease (N = 432) or Ulcerative colitis, (N = 389) or HCs without inflammatory bowel disease (N = 209). Bayesian networks that were generated from this data were used as a structure to inform on the regulatory architecture of MIS-C. This network included CD, UC and HC samples and were constructed using RIMBAnet software(Zhu et al., 2004, 2007, 2008) as previously described.

### Conflicts of interest statement

S.G. reports consultancy and/or advisory roles for Merck, Neon Therapeutics and OncoMed and research funding from Bristol-Myers Squibb, Genentech, Immune Design, Agenus, Janssen R&D, Pfizer, Takeda, and Regeneron.

## Acknowledgements

This work was accomplished by a redeployed workforce supported by the following centers, programs, departments and institutes within the Icahn School of Medicine at Mount Sinai: Mount Sinai COVID-19 Informatics Center; Human Immune Monitoring Center; Program for the Protection of Human Subjects; Department of Psychiatry; Department of Genetics and Genomic Sciences; Department of Medicine; Department of Oncological Sciences; Department of Pediatrics; The Precision Immunology Institute; Tisch Cancer Institute; Icahn Institute for Data Science and Genomic Technology; Friedman Brain Institute; Charles Bronfman Institute of Personalized Medicine; Hasso Plattner Institute for Digital Health; Mindich Child Health and Development Institute; Black Family Stem Cell Institute. S.G. was supported by grants U24 CA224319 and U01 DK124165.

## Author contributions

The following authors contributed to overall study design: AWC, NDB, EES, SGn, BDG, MM, RS. The following authors contributed to establishing the infrastructure for the Mount Sinai COVID-19 Biobank under the purview of which this study was conducted: AWC, NDB, ECh, LL, KMo, NWS, DMDV, GK, EMo, LWi, JLB, CC, RM, SC, AL, LWa, NY, AY, MH, ME, SH, HJ, AA, AT, LEL, LV, BF, AV, The Mount Sinai COVID-19 Biobank Team, CE, SGa, GOA, KAC, KO, KMW, TM, AR, SKS, SGn, BDG, MM. The following authors contributed to experimental design and procedures for RNA sequencing: AWC, NDB, ECh, GEH, NJF, SRT, JS, SB, KMa, EE, EMe, KV, NFi, MT, MS, HX, MP, KA, JH, NK, CB, JL, MY, KT, LS, TM, AR, SKS, SGn, MM, RS. The following authors contributed to data processing and analysis: AWC, NDB, EES, pC, GEH, GB, SRT, HS, YW, SHS, BL, JZ, WW, AK, CA, MEAR, SGn, MM, RS. The following individuals contributed to mining electronic medical records for clinical variables: AWC, NDB, LL, KMo, NWS, DMDV, AV, AK, SJ, RT, ECl, BSG, GN, DB. The following individuals contributed to writing the text and preparing tables and figures for this manuscript: AWC, NDB, EES, PC, ECh, LL, AGB, GEH, NJF, HS, YW, SHS, BSG, GN, SKS, SGn, BDG, MM, RS.

## The Mount Sinai COVID-19 Biobank Team

Charuta Agashe, Priyal Agrawal^1^, Eziwoma Alibo^1^, Kelvin Alvarez^1^, Angelo Amabile^1^, Steven Ascolillo^1^, Rasheed Bailey^1^, Priya Begani^1^, Cansu Cimen Bozkus^1^, Stacey-Ann Brown^1^, Mark Buckup^1^, Larissa Burka^1^, Lena Cambron^1^, Gina Carrara^1^, Serena Chang^1^, Steven T. Chen^1^, Jonathan Chien^1^, Mashkura Chowdhury^1^, Jonathan Chung^1^, Paloma Bravo Correra^1^, Dana Cosgrove^1^, Francesca Cossarini^1^, Arpit Dave^1^, Travis Dawson^1^, Bheesham Dayal^1^, Maxime Dhainaut^1^, Rebecca Dornfeld^1^, Katie Dul^1^, Nissan Eber^1^, Frank Fabris^1^, Jeremiah Faith^1^, Dominique Falci^1^, Susie Feng^1^, Marie Fernandes^1^, Daniel Geanon^1^, Joanna Grabowska^1^, Gavin Gyimesi^1^, Maha Hamdani^1^, Diana Handler^1^, Manon Herbinet^1^, Elva Herrera^1^, Arielle Hochman^1^, Jaime Hook^1^, Laila Horta^1^, Etienne Humblin^1^, Jessica S. Johnson^1^, Subha Karim^1^, Geoffrey Kelly^1^, Jessica Kim^1^, Dannielle Lebovitch^1^, Brian Lee^1^, Grace Lee^1^, Gyu Ho Lee^1^, Jacky Lee^1^, John Leech^1^, Michael B. Leventhal^1^, Katherine Lindblad^1^, Alexandra Livanos^1^, Rosalie Machado^1^, Zafar Mahmood^1^, Kelcey Mar^1^, Glenn Martin^1^, Shrisha Maskey^1^, Paul Matthews^1^, Katherine Meckel^1^, Saurabh Mehandru^1^, Cynthia Mercedes^1^, Dara Meyer^1^, Gurkan Mollaoglu^1^, Sarah Morris^1^, Kai Nie^1^, Marjorie Nisenholtz^1^, Merouane Ounadjela^1^, Vishwendra Patel^1^, Cassandra Pruitt^1^, Shivani Rathi^1^, Jamie Redes^1^, Ivan Reyes-Torres^1^, Alcina Rodrigues^1^, Alfonso Rodriguez^1^, Vladimir Roudko^1^, Evelyn Ruiz^1^, Pearl Scalzo^1^, Alessandra Soares Schanoski^1^, Pedro Silva^1^, Hiyab Stefanos^1^, Meghan Straw^1^, Collin Teague^1^, Bhaskar Upadhyaya^1^, Verena Van Der Heide^1^, Natalie Vaninov^1^, Daniel Wacker^1^, Hadley Walsh^1^, C. Matthias Wilk^1^, Jessica Wilson^1^, Li Xue^1^, Naa-akomaah Yeboah^1^, Sabina Young^1^, Nina Zaks^1^, Renyuan Zha^1^

**Figure S1:** Quality control

A: Scatter plot of normalized expression of sex specific genes show no clear mislabeling. The x-axis is the UTY gene expression (Y chromosome gene) and the y-axis is the XIST gene. The color of the dots represents the sex as reported in the electronic medical records and is defined in the legend. The 1:1 line was added for ease of reading. B: Canonical correlation heatmap of important technical and biological covariates. The heat represents the correlation as defined in the legend and the correlation is shown in each box. The FDR adjusted permutation P-values are shown for significant correlation. C,D: Variance partition violin plot before (C) and after (D) covariate adjustment. X-axes are the different covariates and the y-axes are percent of variation explained by each covariate for each gene.

**Figure S2:** Principal component analyses before (A) and after (B) adjustment for imputed age.

A,B: X and y-axes are the principle components and the amount of variation explained by them. Points are colored by disease states.

**Figure S3:** Cell type deconvolution results.

A,B,C: Box plots showing cell type deconvolution using three references. The x-axes represent deconvoluted cell type as defined in the references, where A and B are from Newman, et al, Nature Biotech, 2019, and C is from Ding, et al, BioRxiv, May 2019. The y-axes are the proportion of the deconvolved expression attributed to each cell type. The color of the box plot represents the disease state as defined in the legend.

D: The correlation heatmap of deconvoluted cell type and measured cell blood count (CBC). The x-axis represents the deconvolution references used for deconvolution, as stated above. The y-axis is the CBC measurements. The color of the heat represents the correlation and adjusted p-values (as defined in Methods) are shown on the plot. Significant adjusted p-values are shown in bold.

**Figure S4:** Module enrichments for cell-type specific signatures.

The x-axis are the modules and the y-axis the cell-types and reference indices: 1= Newman et al, Nature Methods, 2015, 2= Park, et al, Science, 2020, 3= Wilk, et al, Nature Medicine, 2020, 4= Lial, Nature Medicine, 2020, 5= Rowley, et al, Blood, 2011, and 6= Szabo, Nature Communications, 2019. The color and size of the circles represent the log(odds ratio) for the enrichment of the modules for the cell type specific signatures as defined in the legend.

**Figure S5:** Projection of CD8^+^ T-cell subtypes signatures in skyblue and magenta modules.

The x-axis is the name of the signature projected onto the modules as defined in Wherry et al, Immunity, 2007, and the y-axis is the OR for the enrichment of the corresponding signature in the module. The colors of the modules are representative of the module names and are defined in the legend. Enrichment p-values are shown above each bar.

**Data S1:** Clinical Data of MISC and Pediatric COVID-19 cases.

Treatments: treatments received prior to sample collection

MIS-C symptoms

Timeline

COVID-19 Case Summary

**Data S2:** Co-expression Annotations.

Co-expression Network Age Signature Genes Cell Type Signature Enrichments Disease Signature Enrichments GO Term Enrichment MSigDBC7 Term Enrichment MSigDB Hallmark Term Enrichment Skyblue Cell type Dissection

**Data S3:** Celltype Deconvolution.

Celltype Deconvolution Deconvolution Correlation To CBC

**Data S4:** Differential Expression.

Differential Expression Table DE Signature GO Enrichment

**Data S5:**

Module Key Drivers

## Notes

### Competing Interest Statement

The authors have declared no competing interest.

### Funding Statement

This study was funded by the Icahn School of Medicine at Mount Sinai.

### Author Declarations

Institutional Review Board of the Icahn School of Medicine at Mount Sinai

